# Factors associated with vaccine coverage improvements in Senegal between 2005-2019: A quantitative retrospective analysis

**DOI:** 10.1101/2023.03.07.23286913

**Authors:** Hannah K Smalley, Francisco Castillo-Zunino, Pinar Keskinocak, Dima Nazzal, Zoe Sakas, Moussa Sarr, Matthew C. Freeman

**Author notes:** Work performed while the author was with Georgia Institute of Technology prior to joining Amazon (Francisco Castillo-Zunino).

## Abstract

**Objective:** Senegal has demonstrated catalytic improvements in national coverage rates for early childhood vaccination, despite lower development assistance for childhood vaccines in Senegal compared to other low and lower-middle income countries. Understanding factors associated with historical changes in childhood vaccine coverage in Senegal, as well as heterogeneities across its 14 regions, can highlight effective practices that might be adapted to improve vaccine coverage elsewhere.

**Design:** Childhood vaccination coverage rates, demographic information, and health system characteristics were identified from Senegal’s Demographic and Health Surveys (DHS) and Senegal national reports for years 2005 to 2019. Multivariate logistic and linear regression analyses were performed to determine statistical associations of demographic and health system characteristics with respect to childhood vaccination coverage rates.

**Setting:** The 14 administrative regions of Senegal were chosen for analysis.

**Participants:** DHS women’s survey respondents with living children aged 12-23 months for survey years 2005-2019.

**Outcome Measures:** Immunization with the third dose of the diphtheria-tetanus-pertussis vaccine (DTP3), widely used as a proxy for estimating immunization coverage levels and the retention of children in the vaccine program.

**Results:** Factors associated with childhood vaccination coverage include urban residence (β=0·61, p=0·0157), female literacy (β=1·11, p=0·0007), skilled prenatal care (β=1·80, p<0·0001), and self-reported ease of access to care when sick, considering travel distance to a healthcare facility (β=-0·70, p=0·0009) and concerns over traveling alone (β=-1·08, p<0·0001). Higher coverage with less variability over time was reported in urban areas near the capital and the coast (p=0·076), with increased coverage in recent years in more rural and landlocked areas.

**Conclusions:** Childhood vaccination was more likely among children whose mothers had higher literacy, received skilled prenatal care, and had perceived ease of access to care when sick. Overall, vaccination coverage is high in Senegal and disparities in coverage between regions have decreased significantly in recent years.

## Background

Vaccination is one of the most cost-effective and influential public health interventions [1]. For each dollar spent on routine and supplementary vaccinations in low- and middle-income countries from 2011-2020, the average country-level return on investment is estimated to be 44 times the cost [2]. High vaccination coverage is necessary for controlling, eliminating, or eradicating vaccine-preventable diseases [3]. Gavi, The Vaccine Alliance (Gavi), was launched in 2000 and is one of the main sources of vaccine financing for eligible low- and middle-income countries. Some but not all Gavi-supported countries were able to reach the 2015 Global Vaccine Action Plan (GVAP) goal of 90% coverage for the third dose of the diphtheria-tetanus-pertussis vaccine (DTP3) by 2010 [4, 5].

In 2017, Africa reported a DTP3 coverage of 72%, the lowest percentage of all the World Health Organization (WHO) regions [4]. Within the African Region, several countries have outperformed their peers with regards to high and sustained vaccination coverage. DTP3 coverage in Senegal increased from 52% to 93% [6], outperforming other low income (LIC) and lower-middle income (LMIC) countries, despite lower development assistance for health (DAH) relative to many other countries in this group [7, 8],. The Government of Senegal has demonstrated a commitment to improving access to health care, including childhood vaccination, which likely contributed to the country’s success [9-11]. However, like other countries in sub-Saharan Africa, Senegal’s childhood vaccine coverage was not consistent between Senegal’s 14 administrative regions

The purpose of this study was to identify factors associated with DTP3 coverage in Senegal at both the household level and regionally. DTP3 is often used as a proxy for estimating the retention of children in the vaccine program [3]. Subnational trends in early childhood vaccine coverage often illustrate inequities in vaccine service delivery [12, 13], and looking at national-level data alone may overestimate vaccine system performance [14]. Factors statistically associated with full childhood immunization coverage have been identified in previous studies [15-18]. We complement prior work by focusing on DTP3 coverage in Senegal, with an analysis of changes in coverage over time by administrative region from 2005-2019. We analyzed geographic and population factors, including female literacy and access to skilled prenatal care. We also considered factors aggregated at the regional level including poverty and the number of healthcare workers.

## Methods

We conducted a study to assess the demographic associations of early childhood vaccination within Senegal, including assessing the impact of heterogeneities at the subnational level (i.e., between administration regions). For the study period of 2005-2019, we analyzed geographic and population factors for each of Senegal’s 14 administrative regions to examine potential associations with DTP3 coverage. The analysis was conducted as part of the Vaccine Exemplars Program, the purpose of which was to identify how and why some countries have achieved better-than-average and sustained coverage of early childhood vaccines [11, 19-21].

## Data sources

Household-level immunization data was collected from Senegal’s Demographic and Health Surveys (DHS) (years 2005, 2010/2011, 2012/2013, and yearly from 2014 to 2019) [22], with data provided for all 14 regions and all years (notably 3 of the regions were created/split from existing regions in 2008; coverage for these regions in 2005 is estimated to be equal to that of the original combined regions). The data can be accessed upon approval from the DHS Program [23]. DHS surveys were sampled to be representative at the regional level, and thus we were not able to analyze statistical associations at the smaller district level. 28 sampling strata are created which correspond to urban and rural parts for each of the 14 regions; samples are drawn independently from these sampling strata. Specific details on sampling methods for each survey year can be found in the appendices of each report (see [22]). Data extracted from the surveys include DTP3 vaccination (recommended administration to children at 14-weeks-old in Senegal), residence type (urban or rural), female literacy, proportion receiving prenatal care from skilled providers, and self-reported problems accessing care when sick (i.e., women were asked if distance, permission, money, and traveling alone were significant barriers when seeking treatment).

Child immunization is covered in the women’s questionnaire in DHS surveys with responses representing each child of the mother surveyed. Literacy is assessed in the survey by the questioner asking the respondent to read from a card [24]. Prenatal (or antenatal) care is assessed by asking women who attended their last birth, with the following options: doctor, nurse/midwife, auxiliary nurse/midwife, community health worker, other health worker, traditional birth attendant, other, and no antenatal care.

Region-specific population density and poverty (reported for year 2018) were determined from Senegal national reports [25]. Poverty incidence is estimated using the cost of basic needs approach, which considers the proportion of the population unable to meet their basic needs and combines a country-specific food poverty line and non-food poverty line. For more details with respect to this calculation specific to Senegal, see [26]. We collected regional counts of skilled healthcare workers from the Ministère de la Santé et de l’Action sociale (MSAS) (2018) [27].

## Data analysis

Bivariate and multivariate logistic and linear regression analyses were performed to determine statistical associations of factors with respect to coverage of DTP3; factors included in multivariate analysis include residence type (urban or rural), female literacy, self-reported problems accessing care when sick, and prenatal care provider. We calculated DTP3 coverage – the dependent variable in our models - for each region of Senegal using DHS data for each year available during the study period. The immunization coverage for a region was calculated as the percentage of living children ages 12-23 months represented in the survey for that region who received DTP3. Data must be weighted because the overall probability of a household being selected is not constant across regions and residence types [28]. Thus, we compute the individual weight for women and households to adjust for differences in probability of selection and response between cases in a sample. We filter the data for living children ages 12-23 months. For the time series data, we do this for each year of data available. A vaccination is considered administered if it is reported by the mother or marked on the vaccination card with or without a date.

For our analysis, we consider a woman to be literate if she is able to read a whole sentence or part of a sentence as reported in the DHS survey [24, 28]. We consider a woman to have received skilled prenatal care if she answered “doctor”, “nurse/midwife”, or “auxiliary nurse/midwife” as attending her most recent birth.

We calculated the Pearson correlation coefficients (r) to determine statistically significant correlations between the quantitative factors and between factors and DTP3 coverage. We clustered regions geographically into 2 groups (coastal and inland), calculated the variance in DTP3 values over time for each region, and performed independent sample t-tests to compare these two groups of variances.

## Patient and public involvement

There was no patient or public involvement in the study design of this research, the interpretation of the results, or the writing or editing of this document. All data used in this analysis was de-identified public health data.

## Results

Table 1 reports the number of DHS survey respondents (unweighted) in each survey data set which fit the criteria for inclusion (i.e., with living children aged 12-23 months). The number of respondents in each region per year is reported in Table S1 in the supplemental material.

**Table 1.** Senegal household survey data years and unweighted number of survey respondents meeting criteria for inclusion.

| SURVEY YEAR | RESPONDENTS<br>FITTING CRITERIA |
| --- | --- |
| 2005 | 2138 |
| 2010/2011 | 2377 |
| 2012/2013 | 1329 |
| 2014 | 2662 |
| 2015 | 1310 |
| 2016 | 1323 |
| 2017 | 2390 |
| 2018 | 1337 |
| 2019 | 1183 |

### Factors associated with DTP3 coverage by administrative region

For each region in Senegal, Table S2 in the supplemental material reports the weighted percentage of survey respondents with “yes” responses per household factor for year 2019; data is filtered for respondents who meet inclusion criteria. DTP3 coverage by region ranged from 73% to 100% in 2019 (Figure 1, Table S2). Higher DTP3 coverage rates were observed in Dakar (the most populous region and includes the capital city) and its surrounding regions, and along the coast, as well as in the landlocked region of Matam. In 2018, there were approximately 6,600 people per km^2^ in Dakar, which is predominantly urban (97% urban) [29]. The population density in other regions was at most 362 people per km^2^. Population density does not have a statistically significant correlation with DTP3 coverage at the regional level, though this correlation is higher when excluding Dakar (Pearson’s correlation r=0·39, p=0·19).

**Figure 1.**
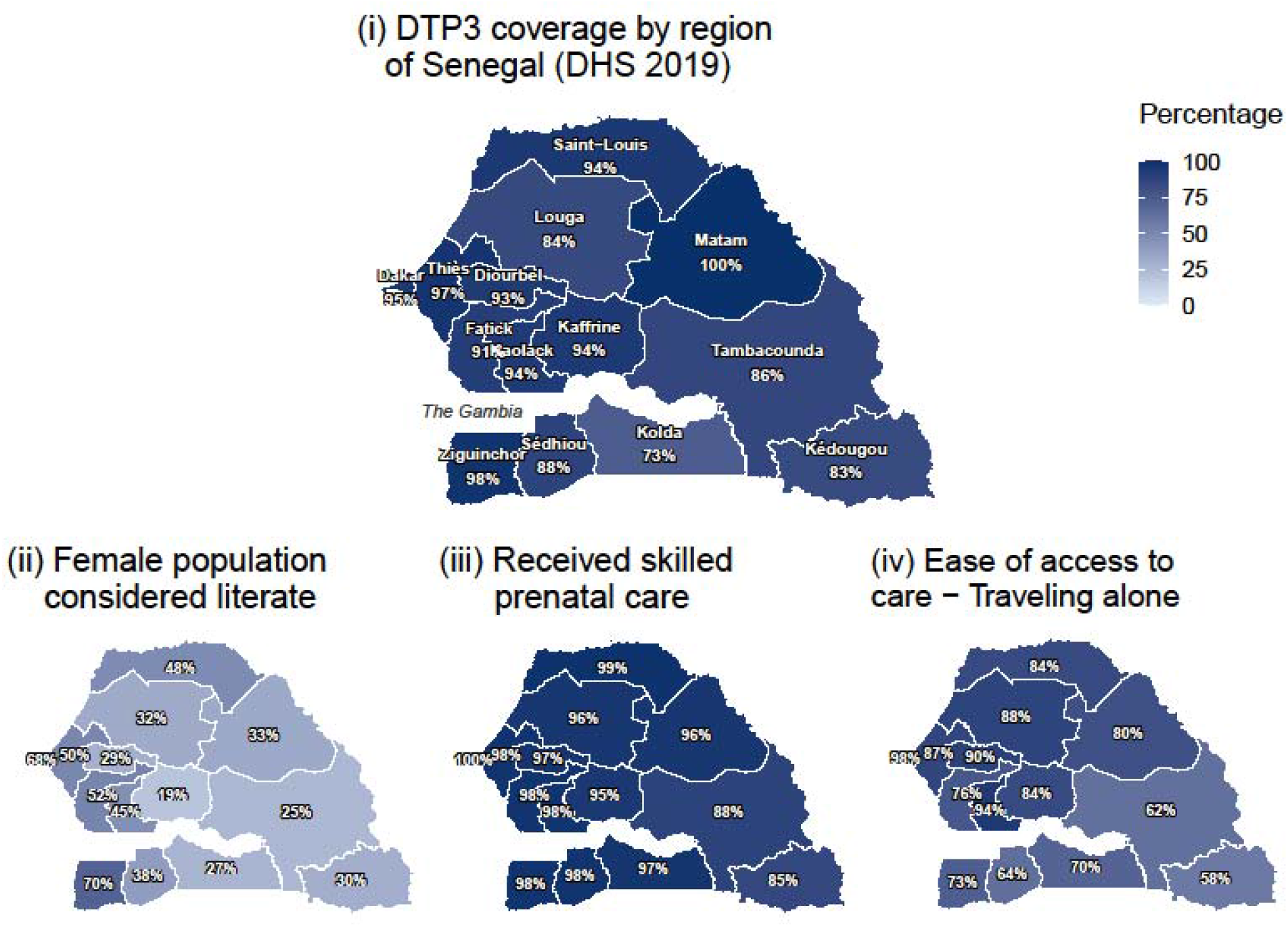
DTP3 coverage by region of Senegal and regional proportions of select metrics [30]. “Skilled Prenatal Care” refers to the proportion of women ages 15-49 who had a live birth within 5 years before the survey who had prenatal care provided by a skilled health care worker (2017). Skilled care includes doctor, nurse, and midwife. Unskilled care includes matron/traditional midwife, “other”, or none. “Ease of Access to Care – Traveling Alone”, refers to the proportion of female survey respondents by region who did not consider traveling alone to be a significant problem with accessing care when sick (2017).

Unadjusted bivariate analyses (Figure 2) revealed significant associations (p=0·05) between higher DTP3 vaccination and the following:

**Figure 2.**
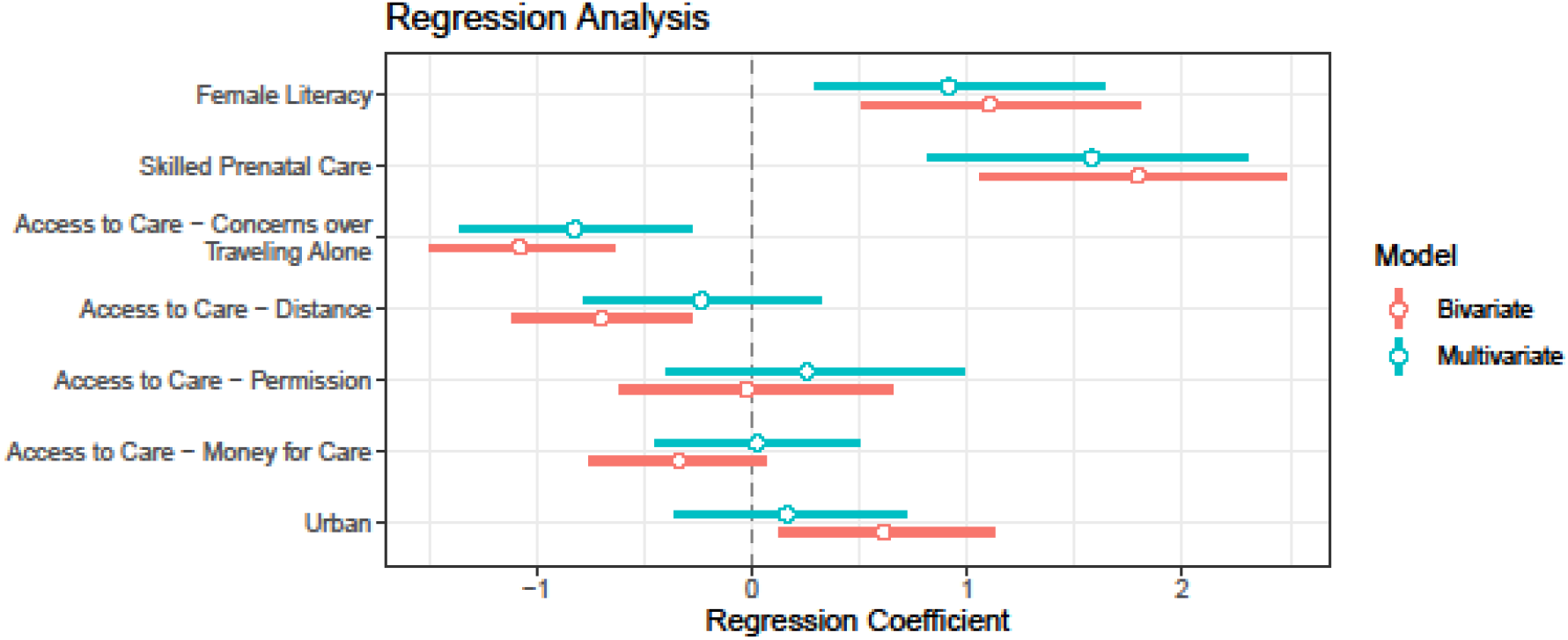
Regression analysis for household-level factors associated with DTP3 vaccination by DHS 2019 survey response per living child aged 12-23 months (weighted N=1129). The bars correspond to 95% confidence intervals. Bars which do not intersect 0 correspond to statistical associations with DTP3 vaccination (< 0 implies negative association, > 0 implies positive association).

- positive associations:
  ○ type of place of residence (urban) (β=0·61, p=0·0157),
  ○ higher female literacy (β=1·11, p=0·0007),
  ○ higher skilled prenatal care (β=1·80, p<0·0001), and
- negative associations:
  ○ more barriers when seeking treatment with respect to
    ▪ distance (β=-0·70, p=0·0009) and
    ▪ traveling alone (β=-1·08, p<0·0001).

We find that significant associations hold for female literacy, skilled prenatal care, distance, and traveling alone when adjusting by type of place of residence (urban).

Multivariate regression revealed correlations between factors (Figure 2), with only 3 factors remaining statistically significant: positive associations with female literacy (β=0·92, p=0·0065) and skilled prenatal care (β=1·58, p<0·0001), and negative associations with more barriers when seeking treatment with respect to traveling alone (β=-0·82, p=0·0026) (more details provided in Table S3 in the supplemental materials). We also found correlations between population density and fewer barriers when seeking treatment with respect to distance (r=0·61, p=0·02) and traveling alone (r=0·48, p=0·08); traveling alone is correlated with distance (r=0·51, p<0·0001).

Coastal regions, namely, Dakar and Ziguinchor, have the highest female literacy rates (68% and 70%, respectively). The percentage of women who received skilled prenatal care from a doctor, nurse, or midwife was lowest in Tambacounda and Kedougou (88% and 85%, respectively), compared to over 95% in other regions (Figure 1).

Poverty level did not statistically explain regional differences in DTP3 coverage (bivariate regression coefficient β=-0·17, p=0·157), potentially due to heterogeneity within regions. We also found no statistical associations between DTP3 coverage and the number of doctors, nurses, and midwives per 100,000 population using bivariate or multivariate regression analysis (Figure 3, more details provided in Table S4 in the supplemental materials).

**Figure 3.**
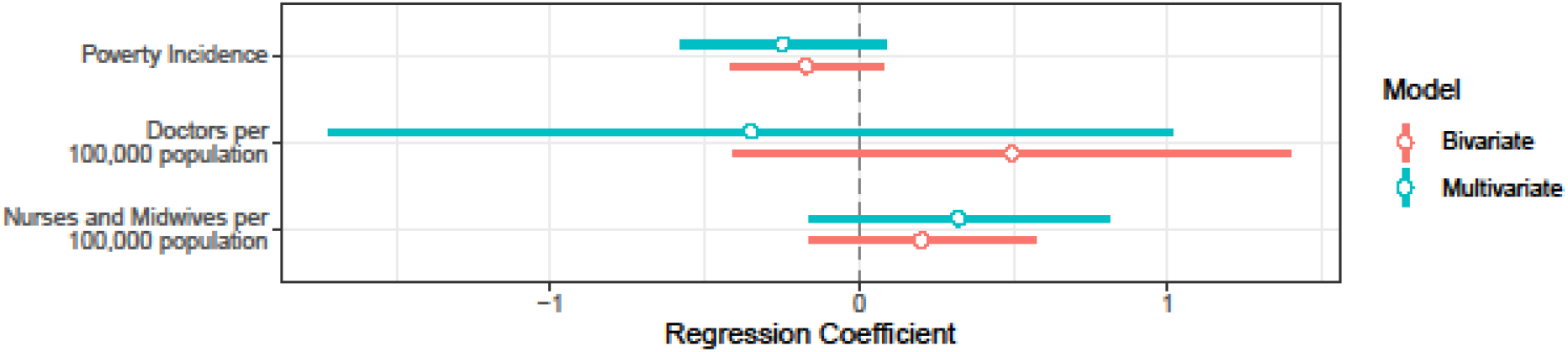
Regression analysis for region-level factor associations with DTP3 vaccination. Poverty incidence is estimated using the cost of basic needs approach, which considers the proportion of the population unable to meet their basic needs and combines a country-specific food poverty line and non-food poverty line [26]. The bars correspond to 95% confidence intervals.

### Access to Health Services

Women in regions with low DTP3 coverage reported more barriers to accessing care with respect to (i) traveling alone to seek care and (ii) distance from healthcare facilities. In the more remote, inland regions of Senegal, as many as 42% of female respondents in 2019 identified traveling alone to seek care as a significant barrier to accessing care. Distance to healthcare facilities was also a substantial barrier to accessing care in these regions of Senegal (identified as a barrier by 50% of female respondents in Tambacounda, the largest region by geographical area). Sociodemographic characteristics of this survey group are reported in Table S5 in the supplemental materials [30].

### Coverage over time by administrative region and geographic location

DTP3 coverage from 2005 to 2019 by region in Senegal is reported in Figure 4 [31]. We observed that the DTP3 coverage was consistently higher for the west and southwestern regions located near Dakar and along the coast, and the variance of DTP3 coverage over time was consistently lower for this group of coastal regions (t-test, p=0.076) compared to the group of eastern and northern regions. The region with the lowest DTP3 coverage in 2019 was Kolda (73%), following a sharp drop in DTP3 coverage after 2017 compared to previous years. The rural and landlocked regions in the north and east demonstrated more variability in coverage over time. However, among these regions, Tambacounda and Kedougou saw large improvements, from 73% to 86% and 60% to 83%, respectively between 2017 and 2019; notably, Matam saw the largest increase (from 73% to 100%) in coverage among all regions between 2010 and 2019.

**Figure 4.**
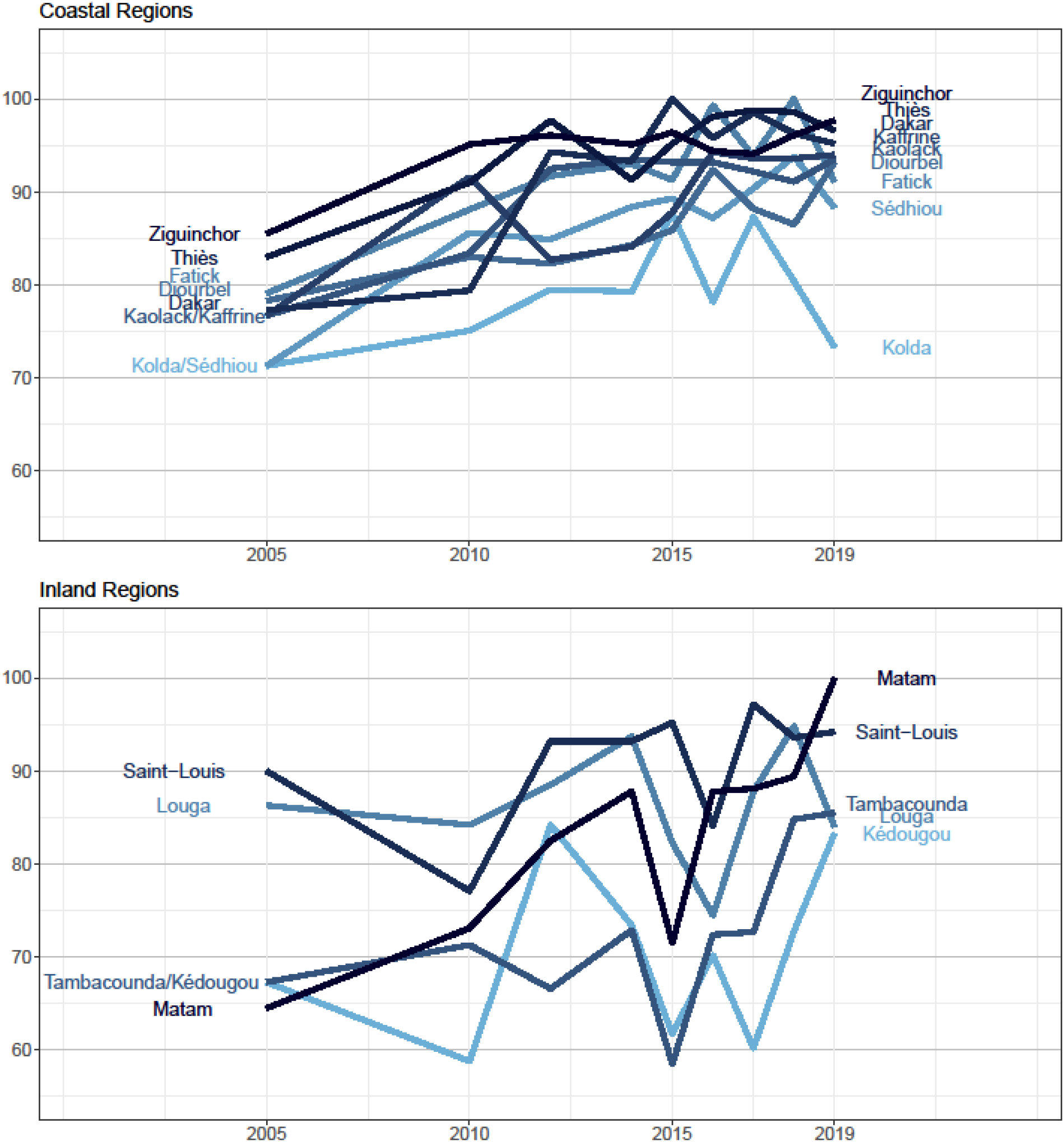
DTP3 coverage in Senegal by year and region (DHS 2005-2019). The regions of Kaffrine, Sedhiou, and Kedougou were split from existing regions in 2008 [31].

## Discussion

We conducted a study to assess the demographic associations of early childhood vaccination within Senegal. Observing heterogeneities in DTP3 coverage across Senegal and comparing those differences with demographic and geographic factors may aid decision-makers in addressing disparities in vaccination access and uptake. Our analysis of household level data revealed associations between DTP3 coverage and 1) female literacy, 2) access to skilled prenatal care, and 3) challenges accessing care while traveling alone.

Early childhood vaccination in Senegal steadily increased for all recommended vaccines from 2002 to 2007, with coverage remaining mostly constant through 2019 [11, 32-37]. Based on an analysis of qualitative data including interviews with Senegalese administrators at various levels and focus group discussions with caregivers and community health workers, Senegal’s immunization program has been successful due to “*strong governance, collaboration, evidence-based decision-making, community ownership, and an overall commitment to health and prioritization of vaccine programming from all stakeholders and government officials*.”[11] Understanding the existing disparities between regions in Senegal, alongside related factors, may support improvements in health equity through utilization of Senegal’s existing governance structures and strong community health worker program.

### Female Literacy & Maternal Education

We found that higher female literacy was significantly associated with higher DTP3 coverage. Female literacy and higher maternal education have been shown to be positively associated with higher vaccination coverage globally [18, 38-40]; it is atypical that Senegal, with low female literacy at the national level [41], has relatively high DTP3 coverage compared to other LMICs. While mothers with high literacy and education may be more likely to independently seek vaccinations for their children [42], the health system in Senegal has been able to reach the more vulnerable rural communities with lower female literacy through community health workers and community-based health huts; “health huts are the backbone of Senegal’s community health system, and they deserve much credit for Senegal’s impressive progress on disease prevention and treatment, including its far-reaching immunization program” [9]. Involvement of community health workers such as respected female elders known as “bajenu gox” have served as a link between the health care services (i.e., health centers, health posts, health huts) and the communities, facilitating and coordinating immunization outreach activities [10].

### Access to Healthcare Workers & Facilities

We found that skilled prenatal care was associated with higher DTP3 coverage in Senegal, which aligns with existing literature. Access to skilled healthcare workers has an important role in childhood vaccination. Limited access to skilled prenatal care has been linked to under-vaccination, or children not receiving all recommended vaccines [15, 43], and a prior study highlighted an association between the number of skilled healthcare workers (nurses and midwives) and DTP3 coverage using a cross-country analysis of developing countries around the world for the years 1990 to 2004 [40]. Self-reported barriers limiting access to care for women when seeking treatment, namely traveling alone to healthcare facilities and travel distance, were associated with DTP3 vaccination. In regions with higher population density, women report fewer concerns regarding traveling alone to access care when sick. Rural communities report more challenges than urban communities with respect to accessing immunization services. Unfortunately, retaining skilled health workers is more difficult in rural regions of Senegal [44]. Senegal has addressed these challenges through training community health workers and health post staff, improving infrastructure for outreach services, increasing capacity for health post expansion, and improving surveillance and data management at local levels [11]. In 2011, a 5-year $40 million grant from USAID helped Senegal construct, maintain, and link more health huts to the national health system [45].

Our results illustrate the role of contextual factors in relation to childhood vaccination. Although national childhood vaccination coverage in Senegal is relatively high compared to other LMICs, there are still gaps in coverage and disparities between regions which, similar to other studies, illustrate inequities in deployment of vaccination services [12, 13]. National health programs rely on the consideration of contextual factors and heterogeneity between subnational areas, such as those identified in this study, to inform programmatic decision-making and improve health equity. Strategies for improving immunization rates need to be tailored considering subnational differences; as stated in The Immunization Agenda 2030, “reaching all people will require higher national vaccination coverage, but also less subnational inequity” [46]. Subnational childhood vaccination has been negatively correlated with institutional mistrust [47]. Strong governance structures are required to prevent and address institutional mistrust while prioritizing equity of healthcare access at the national level [11, 12, 48]. Subnational DTP3 coverage estimates have been compared with national health spending, GDP per capita, and other governance indicators in sub-Saharan Africa to investigate the drivers of equitable immunization services [49]. To reduce differences in vaccination coverage between regions, and address institutional mistrust, national health policies and programs should consider heterogeneity between and within regions. Senegal has implemented top-down and bottom-up mechanisms for decision-making, planning, and resource allocation to address subnational heterogeneity and allow for strategies and initiatives tailored for local populations [11].

### Strengths and limitations

This work uses regionally representative data sets to explain associations between DTP3 coverage and demographic and health system characteristics at the subnational level within Senegal, and evaluates changes in vaccination coverage over time for each of these regions, considering the geographic location of each region. We focused on analyzing respondent groups at the regional level or higher because DHS surveys were not sampled to be representative at the smaller district level. Heterogeneities at the subregional level were thus not evaluated. If district level data were available, we conjecture that negative associations between DTP3 coverage and other factors, such as poverty, could be identified, given heterogeneities at the subregional level. For instances where DTP3 immunization was determined based on maternal recall, errors in recall could have biased results. Additionally, immunization is only reported in the women’s questionnaire of the DHS survey; bias may exist due to families of children with no mothers.

## Conclusions

Senegal has improved vaccination coverage across its 14 administrative regions despite high poverty and low female literacy as compared to other LICs and LMICs [19]. While coverage varies between regions, high coverage has expanded in recent years beyond coastal and urban regions into the inland and rural areas. National and regional efforts - especially increasing the number of health posts and huts to reach rural communities and expanding the community health worker programs - have likely played an important role [9]. Historically, the inland and rural regions have had variability in coverage over time; future years will tell if the recent increases in coverage will be sustained.

## Data Availability

The data that support the findings of this study are available at https://www.dhsprogram.com/data/available-datasets.cfm.

## Acknowledgements

This research was supported in part by the following Georgia Tech benefactors: William W. George, Andrea Laliberte, Joseph C. Mello, Richard “Rick” E. and Charlene Zalesky.

## REFERENCES

1. Rémy V, Zöllner Y, Heckmann U. Vaccination: the cornerstone of an efficient healthcare system. J Mark Access Health Policy. 2015;3:10.3402/jmahp.v3.27041.

2. Ozawa S, Clark S, Portnoy A, Grewal S, Brenzel L, Walker DG. Return On Investment From Childhood Immunization In Low-And Middle-Income Countries, 2011-20. Health Aff (Millwood). 2016;35(2):199-207.

3. World Health Organization. Immunization coverage 2020 2021 [Available from: https://www.who.int/news-room/fact-sheets/detail/immunization-coverage.

4. World Health Organization. Global vaccine action plan 2011-2020 2013 [Available from: https://www.who.int/publications/i/item/global-vaccine-action-plan-2011-2020.

5. World Health Organization. 2018 assessment report of the Global Vaccine Action Plan: strategic advisory group of experts on immunization 2018 [Available from: https://apps.who.int/iris/handle/10665/276967.

6. World Health Organization. WHO vaccine-preventable diseases: monitoring system. 2020 global summary. WHO UNICEF estimates time series for Senegal (SEN) 2020 [Available from: https://apps.who.int/immunization_monitoring/globalsummary/estimates?c=SEN.

7. IHME. Global health spending 1995-2016 2019 [Available from: http://ghdx.healthdata.org/record/ihme-data/global-health-spending-1995-2016

8. WHO and UNICEF. WHO-UNICEF joint reporting form: immunization financing indicators, 2019 [Available from: https://www.who.int/immunization/programmes_systems/financing/data_indicators/en/.

9. Exemplars in Global Health. What did Senegal do? 2021 [Available from: https://www.exemplars.health/topics/under-five-mortality/senegal/what-did-senegal-do.

10. Devlin K, Pandit-Rajani T, Egan KF. Senegal’s Community-based Health System Model: Structure, Strategies, and Learning. Arlington, VA: Advancing Partners & Communities; 2019.

11. Sakas Z, Hester KA, Rodriguez K, Diatta SA, Ellis AS, Gueye DM, et al. Critical success factors for high routine immunization performance: A case study of Senegal. Vaccine: X. 2023;14:100296.

12. Faye CM, Wehrmeister FC, Melesse DY, Mutua MKK, Maïga A, Taylor CM, et al. Large and persistent subnational inequalities in reproductive, maternal, newborn and child health intervention coverage in sub-Saharan Africa. BMJ Global Health. 2020;5(1):e002232.

13. Mosser JF, Gagne-Maynard W, Rao PC, Osgood-Zimmerman A, Fullman N, Graetz N, et al. Mapping diphtheria-pertussis-tetanus vaccine coverage in Africa, 2000–2016: a spatial and temporal modelling study. The Lancet. 2019;393(10183):1843-55.

14. Utazi CE, Thorley J, Alegana VA, Ferrari MJ, Takahashi S, Metcalf CJE, et al. High resolution age-structured mapping of childhood vaccination coverage in low and middle income countries. Vaccine. 2018;36(12):1583–91.

15. Eze P, Agu UJ, Aniebo CL, Agu SA, Lawani LO, Acharya Y. Factors associated with incomplete immunisation in children aged 12–23 months at subnational level, Nigeria: a cross-sectional study. BMJ Open. 2021;11(6):e047445.

16. Sarker AR, Akram R, Ali N, Chowdhury ZI, Sultana M. Coverage and Determinants of Full Immunization: Vaccination Coverage among Senegalese Children. Medicina. 2019;55(8).

17. Ameyaw EK, Kareem YO, Ahinkorah BO, Seidu A-A, Yaya S. Decomposing the rural–urban gap in factors associated with childhood immunisation in sub-Saharan Africa: evidence from surveys in 23 countries. BMJ Global Health. 2021;6(1):e003773.

18. Mbengue MAS, Sarr M, Faye A, Badiane O, Camara FBN, Mboup S, et al. Determinants of complete immunization among senegalese children aged 12-23 months: evidence from the demographic and health survey. BMC Public Health. 2017;17(1):630.

19. Bednarczyk RA, Hester KA, Dixit SM, Ellis AS, Escoffery C, Kilembe W, et al. Exemplars in vaccine delivery protocol: a case-study-based identification and evaluation of critical factors in achieving high and sustained childhood immunisation coverage in selected low-income and lower-middle-income countries. BMJ Open. 2022;12(4):e058321.

20. Hester KA, Sakas Z, Ellis AS, Bose AS, Darwar R, Gautam J, et al. Critical success factors for high routine immunization performance: A case study of Nepal. Vaccine: X. 2022;12:100214.

21. Micek K, Hester KA, Chanda C, Darwar R, Dounebaine B, Ellis AS, et al. Critical success factors for routine immunization performance: A case study of Zambia 2000 to 2018. Vaccine: X. 2022;11:100166.

22. The DHS Program. Senegal Continuous Demographic Health Survey 2005-2019 2019 [Available from: https://www.dhsprogram.com/data/available-datasets.cfm.

23. DHS Program. Demographic and Health Surveys: Data 2023 [Available from: https://www.dhsprogram.com/Data/.

24. The DHS Program. New Data Available from DHS-7 Questionnaire: Literacy, Ownership of Goods, Internet Use, Finances, and Tobacco Use 2017 [Available from: https://blog.dhsprogram.com/dhs7-part-3/.

25. Knoema. Senegal 2021 [Available from: https://knoema.com/atlas/Senegal.

26. Agence Nationale de la Statistique et de la Démographie (ANSD). Deuxieme Enquete De Suivi De La Pauvrete Au Senegal (ESPS-11 2011) 2013 [Available from: https://www.ansd.sn/ressources/publications/Rapport_ESPS-2011.pdf.

27. Ministère de la Santé et de l’Action sociale. Annuaire des statistiques sanitaires et sociales 2018 2020 [Available from: https://sante.sec.gouv.sn/sites/default/files/Annuaire%20Satatistiques%20sanitaires%20et%20sociales%202018.pdf.

28. Croft TN, Marshall AMJ, Allen CK, et al. Guide to DHS Statistics. Rockville, Maryland, USA: ICF.2018.

29. Senegal Data Portal. Population & Financial Statistics of Senegal, 2011 2012 [Available from: https://senegal.opendataforafrica.org/SEPFS2016/population-financial-statistics-of-senegal-2011.

30. Agence Nationale de la Statistique et de la Démographie (ANSD). Sénégal : Enquête Démographique et de Santé Continue (EDS-Continue) 2019 2020 [Available from: https://dhsprogram.com/pubs/pdf/FR368/FR368.T.pdf.

31. Monkam NF. Property Taxation in Senegal: Legislation and Practice. Journal of Property Tax Assessment & Administration. 2011;8(3):41–60.

32. WHO and UNICEF. Senegal: WHO and UNICEF estimates of immunization coverage: 2020 revision 2021 [Available from: https://cdn.who.int/media/docs/default-source/country-profiles/immunization/immunization_sen_2021.pdf.

33. Unicef. Immunization data July 2021 2021 [Available from: https://data.unicef.org/resources/dataset/immunization/.

34. Russo G, Lihui X, McIsaac M, Matsika-Claquin MD, Dhillon I, McPake B, et al. Health workers’ strikes in low-income countries: the available evidence. Bulletin of the World Health Organization. 2019.

35. HRH2030 Program. Three Questions with Dr. Matar Camara: A public health specialist with a vision 2018 [Available from: https://hrh2030program.org/3-questions-with-matar-camara-2/.

36. Despite Senegal’s Nine-Month Health Worker Strike, Nioro Sees LARC Usage Trend Improve: TCI: The Challenge Initiative; 2020 [Available from: https://tciurbanhealth.org/despite-senegals-nine-month-health-worker-strike-nioro-sees-positive-impact-from-larcs/.

37. WHO and UNICEF. Senegal: WHO and UNICEF estimates of immunization coverage: 2019 revision 2020 [Available from: https://www.who.int/immunization/monitoring_surveillance/data/sen.pdf.

38. Streatfield K, Singarimbun M, Diamond I. Maternal Education and Child Immunization. Demography. 1990;27(3):447–55.

39. Vikram K, Vanneman R, Desai S. Linkages between maternal education and childhood immunization in India. Soc Sci Med. 2012;75(2):331–9.

40. Anand S, Bärnighausen T. Health workers and vaccination coverage in developing countries: an econometric analysis. The Lancet. 2007;369(9569):1277-85.

41. The World Bank. Literacy rate, adult female 2020 [Available from: https://data.worldbank.org/indicator/SE.ADT.LITR.FE.ZS

42. Balogun SA, Yusuff HA, Yusuf KQ, Al-Shenqiti AM, Balogun MT, Tettey P. Maternal education and child immunization: the mediating roles of maternal literacy and socioeconomic status. (1937-8688 (Electronic)).

43. Rainey JJ, Watkins M, Ryman TK, Sandhu P, Bo A, Banerjee K. Reasons related to non-vaccination and under-vaccination of children in low and middle income countries: Findings from a systematic review of the published literature, 1999–2009. Vaccine. 2011;29(46):8215-21.

44. Zurn P, Codjia L Fau - Sall FL, Sall Fl Fau - Braichet J-M, Braichet JM. How to recruit and retain health workers in underserved areas: the Senegalese experience. Bull World Health Organ.88(5):386–9.

45. ChildFund. Senegal’s Commitment to Community Health Yields Benefits 2021 [Available from: https://www.childfund.org/senegals-commitment-to-community-health/.

46. World Health Organization. Immunization Agenda 2030: A Global Strategy to Leave No One Behind. World Health Organization; 2020.

47. Stoop N, Hirvonen K, Maystadt J-F. Institutional mistrust and child vaccination coverage in Africa. BMJ Global Health. 2021;6(4):e004595.

48. Shen AK, Fields R, McQuestion M. The future of routine immunization in the developing world: challenges and opportunities. Glob Health Sci Pract. 2014;2(4):381–94.

49. Ikilezi G, Augusto OJ, Sbarra A, Sherr K, Dieleman JL, Lim SS. Determinants of geographical inequalities for DTP3 vaccine coverage in sub-Saharan Africa. Vaccine. 2020;38(18):3447–54.

