## Supplemental Materials for "Factors associated with vaccine coverage improvements in Senegal between 2005-2019: A quantitative retrospective analysis"

| Region | 2005 | 2010/2011 | 2012/2013 | 2014 | 2015 | 2016 | 2017 | 2018 | | 2019 |
| --- | --- | --- | --- | --- | --- | --- | --- | --- | --- | --- |
| Dakar | 150 | 141 | 70 | 130 | 52 | 63 | 164 | 80 | | 60 |
| Diourbel | 229 | 206 | 139 | 259 | 113 | 107 | 182 | 110 | | 107 |
| Fatick | 178 | 185 | 77 | 163 | 94 | 97 | 184 | 90 | | 63 |
| Kaffrine | * | 188 | 118 | 245 | 116 | 140 | 217 | 161 | | 106 |
| Kaolack | 255 | 217 | 131 | 259 | 146 | 124 | 156 | 119 | | 130 |
| Kedougou | * | 87 | 66 | 134 | 78 | 100 | 142 | 76 | | 60 |
| Kolda | 232 | 190 | 108 | 192 | 132 | 104 | 164 | 94 | | 93 |
| Louga | 198 | 165 | 82 | 199 | 97 | 86 | 158 | 92 | | 77 |
| Matam | 178 | 160 | 92 | 182 | 87 | 96 | 180 | 84 | | 94 |
| Saint-Louis | 169 | 157 | 87 | 179 | 59 | 93 | 137 | 79 | | 67 |
| Sedhiou | * | 197 | 95 | 175 | 83 | 78 | 196 | 92 | | 85 |
| Tambacounda | 183 | 178 | 99 | 222 | 91 | 103 | 201 | 106 | | 91 |
| Thies | 232 | 179 | 118 | 214 | 111 | 95 | 190 | 98 | | 107 |
| Ziguinchor | 134 | 127 | 47 | 109 | 51 | 37 | 119 | 56 | | 43 |
| Total | 2138 | 2377 | 1329 | 2662 | 1310 | 1323 | 2390 | 1337 | | 1183 |
| * *Kaffrine, Kedougou, and Sedhiou were split from existing regions in 2008.* | | | | | | | | |  |  |

Table S1 Unweighted number of survey respondents fitting survey criteria (living children ages 12-24 months) by year and region.

| Region | DTP3 | Female Literacy | Skilled Prenatal Care | Access to Care Concerns | | | | Urban |
| --- | --- | --- | --- | --- | --- | --- | --- | --- |
|  |  |  |  | *Traveling Alone* | *Distance* | *Permission* | *Money* |  |
| Dakar | 95.2% | 45.2% | 100.0% | 12.1% | 14.8% | 2.9% | 36.0% | 95.2% |
| Diourbel | 93.2% | 14.3% | 99.3% | 2.8% | 30.0% | 9.2% | 52.7% | 9.7% |
| Fatick | 91.1% | 33.2% | 96.4% | 12.4% | 35.2% | 8.9% | 45.4% | 24.8% |
| Kaffrine | 94.0% | 13.1% | 98.2% | 19.2% | 21.4% | 25.5% | 46.0% | 13.8% |
| Kaolack | 93.5% | 24.8% | 98.4% | 3.7% | 34.7% | 1.3% | 53.4% | 31.2% |
| Kedougou | 83.3% | 24.6% | 89.6% | 21.0% | 7.0% | 19.9% | 32.3% | 20.2% |
| Kolda | 73.3% | 23.7% | 93.5% | 45.2% | 44.5% | 2.0% | 61.3% | 20.7% |
| Louga | 84.0% | 26.3% | 95.9% | 26.5% | 40.8% | 18.6% | 60.4% | 17.9% |
| Matam | 100.0% | 10.3% | 97.4% | 20.5% | 30.9% | 21.3% | 84.3% | 13.8% |
| Saint-Louis | 94.2% | 27.8% | 97.0% | 13.8% | 40.0% | 20.4% | 70.5% | 38.8% |
| Sedhiou | 88.3% | 28.5% | 95.4% | 50.2% | 60.2% | 2.4% | 66.7% | 12.6% |
| Tambacounda | 85.5% | 10.3% | 91.8% | 21.8% | 2.9% | 20.9% | 39.3% | 34.7% |
| Thies | 96.6% | 30.0% | 100.0% | 16.3% | 27.7% | 4.9% | 45.7% | 47.4% |
| Ziguinchor | 97.7% | 61.9% | 97.2% | 14.1% | 16.6% | 2.3% | 43.6% | 40.1% |
| All Senegal | 92.1% | 26.9% | 97.8% | 15.9% | 28.2% | 9.5% | 50.9% | 37.9% |

Table S2 Weighted percentage of survey respondents with “yes” responses per household factor for year 2019. Data is filtered for respondents who meet inclusion criteria.

| **Variable** | **β Coefficients and 95% CI** | **p-value** |
| --- | --- | --- |
| Urban | 0·16 (-0·36, 0·72) | 0·547 |
| **Female Literacy** | **0·92 (0·30, 1·63)** | **0·0065** |
| **Skilled Prenatal Care from a doctor, nurse, or midwife** | **1·58 (0·82, 2·30)** | **< 0·0001** |
| Problems experienced among respondents who sought care when sick: Women ages 15-49 were asked if the following were significant barriers when seeking treatment when they were sick. | | |
| Access to Care - Distance | -0·23 (-0·77, 0·32) | 0·400 |
| Access to Care - Permission | 0·26 (-0·39, 0·98) | 0·463 |
| **Access to Care - Concerns over Traveling Alone** | **-0·82 (-1·35, -0·28)** | **0·0026** |
| Access to Care - Money for Care | 0·02 (-0·45, 0·50) | 0·919 |

Table S3 Household-level factors associated with DTP3 vaccination by survey response per living child age 12-23 months (N=1129 (weighted)): multivariate regression analysis. DHS 2019.

| **Variable** | **Coefficients and 95% CI** | **p-value** |
| --- | --- | --- |
| Poverty Incidence | -0·249 (-0·58, 0·08) | 0·126 |
| Doctors per 100,000 population | -0·352 (-1·72, 1·01) | 0·579 |
| Nurses and Midwives per 100,000 population | 0·323 (-0·16, 0·81) | 0·170 |

Table S4 Region-level factor associations with DTP3 coverage at the regional level (N=14): multivariate linear regression analysis. DHS 2019.

| Sociodemographic Characteristics of Survey Respondents (n=8,649) | |
| --- | --- |
| Religion | |
| Muslim | 97% |
| Christian | 3% |
| Ethnicity | |
| Wolof | 40% |
| Poular | 28% |
| Serer | 15% |
| Mandingue/Soce | 5% |
| Diola | 4% |
| Soninke | 3% |
| Other/non-Senegalese | 5% |
| Education | |
| Primary | 21% |
| Secondary or more | 32% |
| Marital Status | |
| Single | 30% |
| Married | 65% |
| Divorced/separated | 4% |
| Widow | 1% |
| Age | |
| 15-19 | 22% |
| 20-24 | 19% |
| 25-29 | 16% |
| 30-34 | 15% |
| 35-39 | 13% |
| 40-44 | 8% |
| 45-49 | 7% |

Table S5 Sociodemographic characteristics of female survey respondents, 2019.
